# Assessing Long-Lasting Insecticidal Net Coverage, Access, and Utilization: Insights from Malaria-Endemic Regions and Rohingya Camps in Bangladesh

**DOI:** 10.1101/2025.04.17.25326007

**Authors:** Mohammad Sharif Hossain, Amit Kumer Neogi, Ching Swe Phru, Nur-E Naznin Ferdous, Anamul Hasan, Shayla Islam, Md Mushfiqur Rahman, Md Mosiqure Rahaman, Md Nazrul Islam, Shyamol Kumer Das, Abu Toha Md Rezuanul Haque Bhuiyan, Md. Nazmul Islam, Md. Akramul Islam, Mohammad Shafiul Alam

## Abstract

**Introduction:** Malaria remains a major public health challenge, particularly in endemic regions like Bangladesh. To combat this, the National Malaria Elimination Programme (NMEP) has been working to ensure long-lasting insecticidal nets (LLINs) reach vulnerable populations. This study assessed LLIN coverage, access, and utilization among the Bangladeshi population and Forcibly Displaced Myanmar Nationals (FDMN).

**Methods:** A cross-sectional survey was conducted from May to October 2023 across five malaria-endemic districts in Bangladesh and ten FDMN camps in Cox’s Bazar. Data were collected from 1,575 households using structured interviews. Statistical analyses were performed to evaluate LLIN distribution and utilization patterns among different demographic groups, particularly households with pregnant women and under-five children.

**Results:** Among Bangladeshi households, 97.6% owned at least one LLIN, with sufficient coverage for 93.2%. Utilization was high, with 96.4% sleeping under LLINs the previous night. Among pregnant women and under-five children, 95.0% and 98.3%, respectively, used LLINs. However, in FDMN households, while 98.2% owned at least one LLIN, only 44.3% had sufficient coverage, and utilization rates were significantly lower, with 65.7% sleeping under LLINs. Key barriers included inadequate LLIN supply.

**Conclusion:** Bangladesh has made significant progress in LLIN coverage and utilization among its population, surpassing WHO’s 80% threshold. However, gaps remain in the FDMN population, necessitating targeted interventions to achieve universal coverage and further reduce malaria morbidity and mortality.

## Introduction

Among the mosquito-borne diseases, malaria is the oldest one that causes many clinical symptoms such as a sensation of cold, fever, chills, headaches, nausea, vomiting, sweating, joint pain, prostration, and malaise (1). The World Malaria Report 2023 claimed an estimated 249 million malaria cases in 2022 in 84 malaria endemic countries which was a 5 million increase from 2021. Apart from the cases, the estimated number of deaths decreased from 631, 000 in 2019 to 608,000 in 2022.However, the percentage of total malaria deaths in children under five years has shown no change since 2015 (2).

Bangladesh has developed a National Strategic Plan (NSP) for Malaria Elimination 2024-2030 that targets a malaria-free country by 2030, aligned with the “Global Technical Strategy for Malaria 2016-2030”. The goal of this plan is to, by 2027, attain zero mortality due to indigenous malaria and maintain this status and, by 2030, interrupt local transmission of indigenous malaria in a phased manner and prevent the re-establishment of local transmission (3). Among 64 districts, only 13 north-eastern and south-eastern districts of Bangladesh bordering India and/or Myanmar are malaria endemic(4). An estimated 17.7 million people are at risk of malaria in those endemic areas. Based on the annual malaria incidence, three districts of the Chattogram hill tracts (CHT): Khagrachari, Rangamati, and Bandarban are considered as the hyper-endemic areas as they contributed to the majority of the cases (3). *Plasmodium falciparum* (*Pf*) is the most common malaria parasite in Bangladesh followed by *Plasmodium vivax (Pv).* The other two species, *Plasmodium malariae (Pm)* and *Plasmodium ovale (Po),* are rare. In 2023, the total number of malaria cases was reduced to 16,567 from 18,195 and deaths increased from 6 to 14 compared to 2022 (5). Of the total cases in 2023, only *Pv* alone accounted for 45.3% and this increase has been rapid like 5% in 2011 to 20.3% in 2020 and in 2022, it was 32.1% (6, 7).

In 1978, Myanmar imposed the Emergency Immigration Act under the military regime on the minority Muslim Rohingyas which started the Rohingya crisis and led to the Muslim Rohingyas flocking to Bangladesh (8). In 2011, the United Nations Refugee Agency (UNHCR) reported that around 265,000 Rohingya were residing in Bangladesh and of them, 200,000 were undocumented by the Government of Bangladesh (GoB) and other non-governmental organizations (NGOs) (9). On August 25, 2017, Myanmar started an exodus in Rakhine State and as a result, more than 712,179 sought asylum in Bangladesh (10). By October 2019, an estimated 911,566 Rohingya refugees were seeking asylum in the Cox’s Bazar district of Bangladesh (11). Almost all of them reside in 34 camps of Cox’s Bazar which is also a malaria endemic area. In 2022, a total of 65 malaria cases were detected and it rose to 94 in 2023, and between January and June of 2024, the number reached 244.

The National Malaria Elimination Programme (NMEP), together with the NGO consortium led by BRAC, has achieved remarkable success in malaria control through continuous support from the Global Funds to Fight AIDS, Tuberculosis and Malaria (GFATM) and the GoB in 13 malaria endemic districts since 2008 (12). Among the goals, two of them were: 1) to provide long-lasting insecticidal nets (LLINs) to 100% of households in the three malaria-endemic districts with the highest malaria burden and 80% coverage in the other ten malaria-endemic districts; 2) to provide periodic (every 3 years) treatment of non-LLINs with suitable insecticides (12).Between July 2021 and June 2024, a total of 350,000 LLINs was distributed among the Forcibly Displaced Myanmar Nationals (FDMN) population at-risk through mass campaigns.

BRAC and icddr,b jointly conducted the first-ever malaria prevalence survey of Bangladesh in 2007 (13) followed by a follow-up survey in 2013 supported by NMEP (4). From these surveys, it was found that the use of the– insecticidal bed-nets significantly increased in 2013 compared with 2007. Since 2007, the BRAC-led consortium of 20 smaller partner NGOs has been working on a malaria control and prevention programme through insecticidal bed-net distribution, early diagnosis and management, providing treatment according to the national guideline, referral of complicated cases to the nearest district hospital for better management, and Information, Education & Communication (IEC) activities. Through a survey, it was also found that the net utilization percentage among pregnant women and children under five was 85% and 90%, respectively (14). However, to keep track of the program’s performance, the survey of the use of insecticidal bed-nets (LLIN/ITNs) needs to be done annually.

To track the success of the program, from 2008 and onward, a regular survey has been conducted to estimate the utilization of LLINs in malaria-endemic areas of Bangladesh. On the other hand, in 2021, a small study was conducted to assess the coverage and utilization of LLINs among the FDMN. The study found very poor coverage and utilization in those areas (15). The objective of this study is to assess the coverage, access, and utilization of LLINs among the Bangladeshi people and to understand the perception of the Rohingya people regarding the LLIN use in FDMN camp areas.

## Materials and Methods

### Study Design

Aligned with the study objectives, this cross-sectional study will measure the coverage of LLINs which are NMEP’s concern about how people in the malaria endemic areas are accepting LLIN in their daily practice, whether the recipient households are using and caring for this in the appropriate manner, especially for the ethnic minorities, pregnant women, under-5 children in CHT, Non-CHT and FDMN areas.

### Study Area

The cross-sectional survey unfolded in two phases: 1) encompassing five malaria-endemic districts of Bangladesh, consider five malaria endemic districts and distribution strategy of LLIN coverage and 2) spanning 10 Rohingya camps in FDMN, Cox’s Bazar. Of the five selected districts, three are hyper-endemic districts (Khagrachari, Rangamati, and Bandarban), and the other two are low-malaria-endemic districts (Cox’s Bazar and Chattogram). We randomly selected three upazilas from each of the Khagrachari, Rangamati, and Bandarban districts and two upazilas from each of the Cox’s Bazar and Chattogram districts. Thus, in total, 13 upazilas were included in this study. From each upazila, three villages were selected randomly from the list of the villages where LLINs had been distributed by NMEP to households with children under 5 years of age or pregnant women, resulting in the final selection of 39 villages for data collection **(Fig 1).**

**Fig 1.**
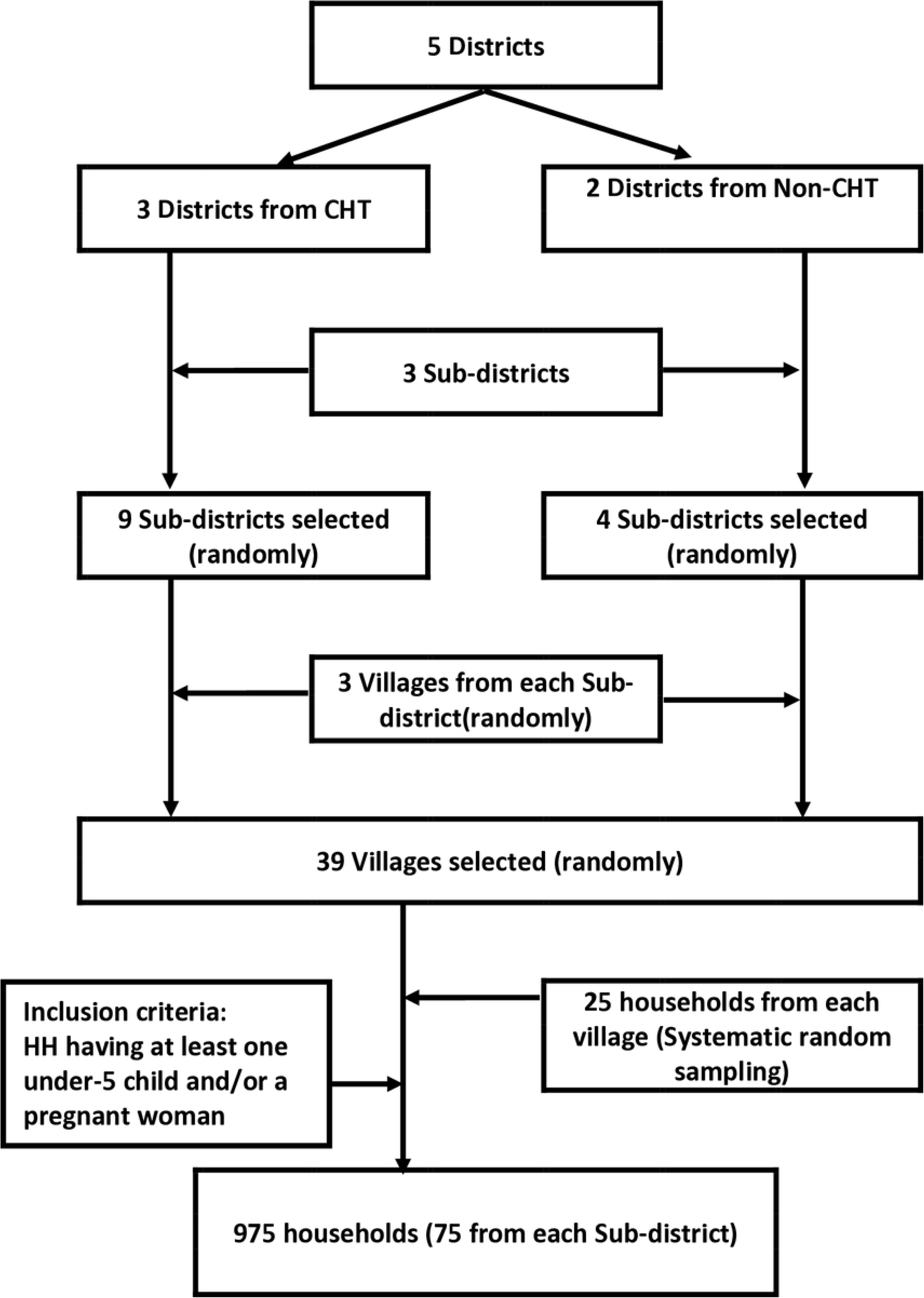
Study flow chart for sampling technique among Bangladeshi nationalities.

In FDMN areas, the LLINs were distributed across 34 camps at the onset of the crisis in 2018. From these 34 camps, we randomly selected 10 for the LLIN cross-sectional survey. Again, in a village or camp, data were collected from households during a single visit. During that visit, we asked the head or an adult member of the selected household whether they were interested in participating in this study. If interested, they were asked to provide the written informed consent and then were enrolled in the study.

### Sample Size

Previous study found that (15), the overall 90.9% (CHT: 99.3% and Non-CHT: 72.0%) of households in Bangladeshi community had LLINs. Thus, we expected that at least 95% of households would be using LLINs. Assuming the difference between the estimated and true prevalence (i.e. the design effect of 1.5% and 5% statistical level of significance), a minimum total of 811 samples would be required for this study. Considering a 10% refusal rate to provide the information, at least a total of 893 households needed to be surveyed. Information was collected from 25 households in each village, and therefore, a total sample of 75×39=975 households was included in the study.

In FDMN areas, a study was conducted on a smaller scale in 2021 (15) and found that the household ownership of having LLINs was very low, only 68%. Thus, by considering that 70% of households were using LLINs and assuming a design effect 4% and a 5% statistical level of significance, minimum total of 505 samples would be required for this study. Considering a 10% refusal as well, a total of 556 households needed to be surveyed. A total of 600 households (on average 60 households from each camp) were interviewed from these 10 camps through the Probability proportional to size (PPS) sampling method **(Table 1)**.

**Table 1.** FDMN population study area for LLIN survey.

| Upazila | Name of the areas | Number of camps | Total HH | Targeted HH | HH surveyed |
| --- | --- | --- | --- | --- | --- |
| Ukhiya | Kutupalong | 2 extension | 6650 | 67 | 67 |
|  |  | KRC | 3242 | 32 | 32 |
|  |  | 4 | 8194 | 82 | 82 |
|  |  | 7 | 8345 | 84 | 84 |
|  | Balukhali | 9 | 7808 | 78 | 78 |
|  | Palongkhali | 15 | 13750 | 138 | 138 |
|  |  | 21 | 3614 | 24 | 24 |
| Teknaf | Leda | 24 | 7616 | 47 | 47 |
|  |  | 27 | 3579 | 33 | 33 |
|  |  | NRC | 3705 | 15 | 15 |

HH, Household; KRC, Kutupalong Registered Camp; NRC, Nayapara Registered Camp.

### Data collection tools and techniques

Data were collected between May and October 2023 through face-to-face interviews with the household heads or the representatives of the household using a pre-tested structured questionnaire (**S1 File**). Besides socio-demographic and household economy data, the questionnaire included information on the knowledge and practice of the household on different aspects of malaria, availability and use of LLINs especially by vulnerable groups (e.g., under-five children and pregnant women), knowledge of means of using insecticidal bed nets, and compliance with treatment. Data were collected in tabs and transferred in real-time to the online server whenever available.

### Statistical analysis

Data entry into the tabs was conducted using Open Data Kit (ODK) software version 2021.4.0 on the Android platform. To ensure data quality, a pre-designed data-check program implemented in STATA was utilized to identify inconsistencies. Statistical analyses were performed using the Stata software, version 15.1 (Stata Corporation, College Station, Texas, USA).

Baseline characteristics for two groups, CHT and Non-CHT, were analyzed using summary statistics, including the number, mean, median, inter-quartile range, standard deviation, minimum, and maximum. Unpaired t-tests were employed to compare quantitative variables between the two groups, while categorical variables were assessed using either the Chi-square test or Fisher’s exact test. Proportions between groups were compared using Z-statistics. A significance level of *p*<0.05 was considered statistically significant in all analyses.

### Informed consent

To ensure accurate translation of the consent forms into the local language, a comprehensive forward-and-backward translation process was implemented. However, in FDMN areas, we adhered to both Bangla and English version of the consent form in accordance with the regulations of the Refugee Relief and Repatriation Commissioner (RRRC). Before obtaining any study-related information, the written informed consent was secured from all participants in presence of local interpreter, known as “Majhi”.

The consent process involved providing participants with detailed information about the study’s objectives, the nature of the data collected, potential benefits and risks, and a guarantee of confidentiality for all information and results generated by the study. The information and consent form were verbally communicated to all participants, with illiterate individuals expressing their consent through a fingerprint in the presence of a witness.

Participants were explicitly informed of their right to withdraw their consent at any point during the interview without the obligation to provide a reason or fear any negative consequences.

### Ethical approval

This study was conducted after obtaining formal approval (PR-20097) from the Ethical Review Committee of icddr,b, Mohakhali, Dhaka. No participants were interviewed without obtaining prior informed written consent. The study information linked to participant identification was kept strictly confidential and inaccessible without due permissions.

### Result

A total of 1,575household were interviewed and among them 61.9% were from Bangladeshi population and 38.1% were from FDMN **(Table 2)**. Among the Bangladeshi respondents, only 13.9% were household head whereas 44.8% were in FDMN. Of the Bangladeshi respondents, 85.0% were female,and the median age was 28 years (inter-quartile range: 24 – 35 years), but among the FDMN, 66.5% were female, and the median age was 32 years (inter-quartile range: 26–42 years). A majority of FDMN household heads (63.5%) were unemployed, while the highest portion of Bangladeshi household heads were daily laborer (24.9%).

**Table 2.** Demographic Characteristics of the study population.

| Characteristics | Bangladeshi respondents,<br>N=975; n (%) | FDMN respondents,<br>N=600; n (%) |
| --- | --- | --- |
| **Type of the respondent** |  |  |
| Household head | 135 (13.9) | 269 (44.8) |
| Spouse of household head | 686 (70.4) | 276 (46.0) |
| Other | 154 (15.8) | 55 (9.2) |
| **Sex** |  |  |
| Male | 146 (15.0) | 201 (33.5) |
| Female | 829 (85.0) | 399 (66.5) |
| **Age, year** |  |  |
| Median (IQR) | 28 (24 - 35) | 32 (26 - 42) |

| **Education** |  |  |
| --- | --- | --- |
| No schooling | 193 (19.8) | 414 (69.0) |
| 1 - 5 Years | 302 (31.0) | 126 (21.0) |
| 6 - 9 Years | 283 (29.0) | 41 (6.8) |
| 10-12 Years | 178 (18.3) | 12 (2.0) |
| Graduate | 14 (1.4) | 4 (0.7) |
| Other | 5 (0.5) | 3 (0.5) |
| **Occupation of Household Head** |  |  |
| Agriculture | 188 (19.3) | 0 (0.0) |
| Jhum Cultivation/Rubber Plantation | 79 (8.1) | 0 (0.0) |
| Unemployed | 0 (0.0) | 381 (63.5) |
| Own business | 150 (15.4) | 26 (4.3) |
| Rickshaw/Van/Car/Boat driver/ | 144 (14.8) | 13 (2.2) |
| Carpenter/Mason | 243 (24.9) | 119 (19.8) |
| Daily labour | 107 (11.0) | 51 (8.5) |
| Service other | 64 (6.6) | 10 (1.7) |
| **Family Size** |  |  |
| ≤ 4 | 353 (36.2) | 190 (31.7) |
| ≥ 5 | 622 (63.8) | 410 (68.3) |

IQR, Interquartile range.

Regarding the LLIN coverage, in the CHT region, a small percentage of participants (1.6%) had no LLIN, while the majority had 3-4 bed-nets (48.3%), while in the Non-CHT region, a slightly larger proportion (4.0%) had no LLIN, with the majority having 55.6% with 1-2 bed-nets. Regarding the sufficiency of LLINs, in the CHT regions, 95.5% of households had sufficient LLINs, and in Non-CHT regions, it was 87.9% of LLINs. However, 98.2% of FDMN households had LLINs, with 93.0% having 1-2 bed-nets, but the sufficient number of LLINs was only 44.3% **(Table 3)**.

**Table 3:** Coverage of LLINs in the study areas of Bangladesh.

| Characteristics | Study Area |  |  |  |
| --- | --- | --- | --- | --- |
|  | Bangladeshi |  |  | FDMN |
|  | CHT (n=675) | Non-CHT (n=300) | Total (n=975) | Total (n=600) |
| **Household have any LLIN?** |  |  |  |  |
| Yes | 664 (98.4) | 288 (96.0) | 952 (97.6) | 589 (98.2) |
| No | 11 (1.6) | 12 (4.0) | 23 (2.4) | 11 (1.8) |
| **How many LLIN? (among household have LLIN)** |  |  |  |  |
| 1-2 Bed-nets | 271 (40.1) | 160 (55.6) | 431 (45.3) | 548 (93.0) |
| 3-4 Bed-nets | 321 (48.3) | 105 (36.5) | 426 (44.8) | 39 (6.6) |
| 5 or more Bed-nets | 72 (10.8) | 23 (7.9) | 95 (9.9) | 2 (0.3) |
| **Household have sufficient LLIN? (among household have LLIN)** |  |  |  |  |
| Yes | 634 (95.5) | 253 (87.9) | 887 (93.2) | 261 (44.3) |
| No | 30 (4.5) | 35 (12.1) | 65 (6.8) | 328 (55.7) |
| **How many LLIN are usable now (not ripped/ruined)? (among household have LLIN)** |  |  |  |  |
| None | 28 (4.2) | 25 (8.7) | 53 (5.6) | 116 (19.7) |
| 1-2 Bed-nets | 292 (44.0) | 172 (59.7) | 464 (48.7) | 455 (77.2) |
| 3-4 Bed-nets | 300 (45.2) | 76 (26.4) | 376 (39.5) | 18 (3.1) |
| 5 or more Bed-nets | 44 (6.6) | 15 (5.2) | 59 (6.2) | 0 (0.0) |
| **Where did you get the LLINs from?** |  |  |  |  |
| Global Fund | 664 (100.0) | 288 (100.0) | 952 (100.0) | 322 (56.4) |
| Another Source | 0 (0.0) | 0 (0.0) | 0 (0.0) | 64 (10.9) |
| Don't Know | 0 (0.0) | 0 (0.0) | 0 (0.0) | 193 (32.8) |

The findings from this survey indicate that, on average, there was approximately one LLIN for every 2 people (2696 LLINs for 5249 persons), and among the FDMN population, it was for every 3.9 person (875 LLINs for 3,419 persons). In CHT areas, the proportion of population per LLIN was 1.8 (1944 LLINs for 3538 persons) which is the target coverage by the WHO. In contrast, in Non-CHT areas, this proportion was 2.3 (752 LLINs for 1711 persons) (**Fig 2**).

**Fig 2.**
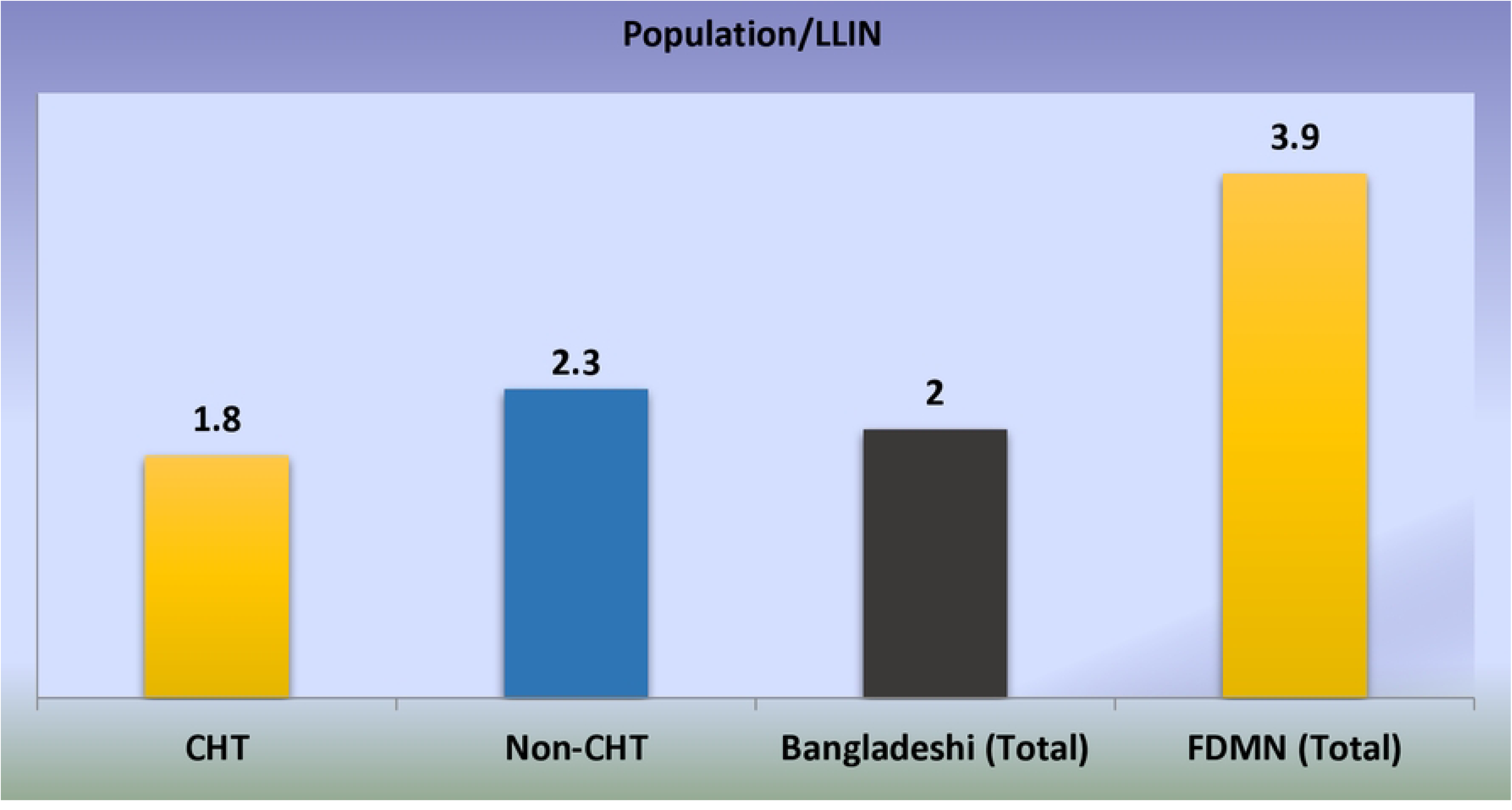
Rate of population per LLIN in the study area.

Overall, more than 93% of Bangladeshi respondents indicated that all of their household members were able to sleep under LLINs; however, it was 93.1% and 92.4% among households with under-five-year-old children and pregnant women, respectively. In contrast, in FDMN, 41.4% reported that all of their household members were able to sleep under the LLINs, and among households with under-five-year-old children and pregnant women, only 45.3% and 43.0%, respectively were able to do so. While asking the reasons why all household members can’t sleep under the LLINs, the primary reason cited in both the regions was “Inadequate number of LLINs” (98.5% in Bangladeshi population and 86.7% in FDMN. The primary reasons cited for household members sleeping under the LLIN every night in both regions was “Fear of mosquito bites” (66.6% in Bangladeshi population and 91.8% in FDMN).When asked the reason why household members do not sleep under the LLIN every night in both the Bangladeshi(98.8%) and FDMN(86.9%) regions, the primary reason was “Inadequate number of LLINs” **(Table 4)**.

**Table 4.** Accessibility of LLINs in the household in the study area.

| Characteristics | Study Area |  |  |  |
| --- | --- | --- | --- | --- |
|  | Bangladeshi |  |  | FDMN |
|  | CHT (n=664) | Non-CHT (n=288) | Total (n=952) | Total (n=600) |
| **All the household member can sleep under the LLIN?** |  |  |  |  |
| Yes | 634 (95.5) | 253 (87.9) | 887 (93.2) | 244 (41.4) |
| No | 30 (4.5) | 35 (12.1) | 65 (6.8) | 345 (58.6) |
| **All the household (household had under 5 years children) member can sleep under the LLIN?** |  |  |  |  |
| Yes | 594 (95.7) | 235 (87.4) | 829 (93.1) | 185 (45.3) |
| No | 27 (4.3) | 34 (12.6) | 61 (6.9) | 223 (54.7) |
| **All the household (household had pregnant woman) member can sleep under the LLIN?** |  |  |  |  |
| Yes | 59 (95.2) | 38 (88.4) | 97 (92.4) | 37 (43.0) |
| No | 3 (4.8) | 5 (11.6) | 8 (7.6) | 49 (57.0) |
| **Why all the household member can't sleep under the LLIN?** |  |  |  |  |
| Inadequate no. of LLIN | 30 (100.0) | 34 (97.1) | 64 (98.5) | 299 (86.7) |
| LLIN is ripped | 0 (0.0) | 1 (2.9) | 1 (1.5) | 36 (10.4) |
| Other | 0 (0.0) | 0 (0.0) | 0 (0.0) | 10 (2.9) |
| **Does every member of your Household sleep under LLIN every night?** |  |  |  |  |
| Yes | 624 (94.0) | 247 (85.8) | 871 (91.5) | 243 (41.3) |
| No | 40 (6.0) | 41 (14.2) | 81 (8.5) | 343 (58.2) |
| Don't know | (0.0) | (0.0) | (0.0) | 3 (0.5) |
| **Reason of all the household member sleep every night under the LLIN** |  |  |  |  |
| Fear of malaria | 105 (16.8) | 53 (21.5) | 158 (18.1) | 12 (4.9) |
| Fear of mosquito bits | 420 (67.3) | 160 (64.8) | 580 (66.6) | 223 (91.8) |
| To stay free of disease | 99 (15.9) | 34 (13.8) | 133 (15.3) | 6 (2.5) |
| Other | 0 (0.0) | 0 (0.0) | 0 (0.0) | 2 (0.8) |
| **Reason of all the household member do not sleep every night under the LLIN** |  |  |  |  |
| Inadequate number of LLIN | 40 (100.0) | 40 (97.6) | 80 (98.8) | 298 (86.9) |
| LLIN is ripped | 0 (0.0) | 1 (2.4) | 1 (1.2) | 33 (9.6) |
| Other | 0 (0.0) | 0 (0.0) | 0 (0.0) | 12 (3.5) |

Of the total population, 96.4% slept under LLINs the night before the survey, whereas in FDMN, it was only 65.7%. When comparing the study area (CHT) with non-CHT areas, 97.3% and 94.4% of participants, respectively, used LLINs in the CHT and non-CHT areas. The difference between the two areas is statistically significant (*p*-value < 0.001). Among the pregnant women and children under the age of five, 95.0% and 98.3%, respectively, slept under LLINs the previous night of the survey, while in FDMN, this percentage was 77.4% and 78.3%, respectively (**Table 5**).

**Table 5.** Utilization of LLINs the previous night of the survey in study area.

| Characteristics* | Study Area |  |  |  |
| --- | --- | --- | --- | --- |
|  | Bangladeshi |  |  | FDMN |
|  | CHT<br>% (n/N) | Non-CHT<br>% (n/N) | Total<br>% (n/N) | Total<br>% (n/N) |
| % total population who slept under LLINs at the previous night | 97.3<br>3263/3353 | 94.4<br>1489/1577 | 96.4<br>4752/4930 | 65.7<br>2126/3234 |
| % under-five children who slept under at the previous night | 98.0<br>697/711 | 98.8<br>318/322 | 98.3<br>1015/1033 | 78.3<br>474/605 |
| % pregnant women who slept under LLINs at the previous night | 95.0<br>57/60 | 95.1<br>39/41 | 95.0<br>96/101 | 77.4<br>65/84 |
| % male who slept under LLINs at the previous night | 97.6<br>1573/1611 | 93.6<br>716/765 | 96.3<br>2289/2376 | 64.3<br>982/1526 |
| % female who slept under LLINs at the previous night | 97.0<br>1690/1742 | 95.2<br>773/812 | 96.4<br>2463/2554 | 67.0<br>1144/1708 |

\* # of individual slept last night under LLIN / # of individual present at the household last night

## Discussion

The indicators of LLIN coverage in the high and low endemic area of Bangladesh and in the area of FDMN were investigated in this study. In total, 97.6% of the households among the Bangladeshi population and 98.2% had at least one LLIN in their household. Compared to the previous study among the Bangladeshi population in this region, reported LLIN coverage is significantly higher (16) but almost similar to another study (17), indicating ongoing efforts by the government and other NGOs to combat malaria realigned with the national strategy to ensure that 100% of households in malaria hyper-endemic districts receive LLINs, while also implementing targeted LLIN coverage in other malaria-endemic districts(12).The proportion of LLIN/ITN coverage in both areas is similar to the neighboring country India(18)and Myanmar (19), but higher than in Nepal (20) and some African countries (21–23). As recommended by the WHO, the objective is to guarantee one LLIN for every two household members in order to attain universal coverage (24), and the NMEP of Bangladesh has been very successful with the help of local NGOs lead by BRAC. The present study revealed that there was a supply of at least one LLIN for every 1.8 household members in the malaria hyper-endemic areas. However, the general coverage was one for every two household members and these findings were similar to Ethiopia (25) and Kenya (26); nevertheless, the FDMN population has not yet met this benchmark.

The availability of LLINs was noted to be exceptional. Among the households that received LLINs, 93.2% reported that every family member was able to sleep under the LLIN each night. The fact that so many households in the area owned LLINs shows how important mass-distribution efforts were for making sure that people who lived in places where malaria was common own LLINs. This access rate was much higher compared with the study conducted in Thailand (86.1%) (27), Ethiopia (88.9%) (28) and Nigeria (33.0%) (29). The proportion of all the family member could be able to sleep under the LLINs of the household having at least one under five years of children was 98.3% and among households having at least one pregnant woman, it was 95.0%. While in FDMN, the percentage was very low (41.4%).However, among the household having at least one child under five years of age and least one pregnant woman, the proportions were close to the levels recommended by the WHO (30).

The present study revealed that among the Bangladeshi population, 96.4% of total population slept under LLIN the night prior to the interview which was much higher compared with previous study (13), and much higher than the WHO’s recommended optimal threshold (80%) (31). The utilization rate was superior in comparison to other malaria-endemic regions such as India (59.4%) (18), Nigeria (86.0%) (32), and Ethiopia (38.4%) (28). It is crucial to ensure that the most vulnerable population are using ITN/LLINs extensively and regularly in order to control morbidity and mortality due to malaria, particularly in high per-endemic areas. This study also revealed that 95.0% Bangladeshi pregnant women slept under the LLINs in previous night of interview, which was much higher compared with other malaria-endemic regions such as Burkina Faso (57.6%) (33), Madagascar (68.5%) (23), Ethiopia (79.1%) (34), Cameroon (82.5%) (35) and the bordering country India (89.0%) (36). This rate among the children under five years of age in this study among the Bangladeshi population was 98.3%. The utilization of LLINs among children under five in this area was comparable to that in India (96.0%) (36), but significantly greater than in other malaria-endemic countries such as Madagascar (80.8%) (23), Nigeria (80.0%) (32), Liberia (39%) (37), and Kenya (59.0%) (38). Moreover, the overall utilization in FDMN was fairly low (65.7%), but higher compared with India (18) and Ethiopia (28). The proportions of utilization among the pregnant women (77.4%) and children under five years of age (78.3%) were also low. The sleeping pattern between males (96.3%) and females (96.4%) was high and fairly similar among the Bangladeshi population, which was much higher than in India (18) and Rwanda (39).

## Limitations

The limitations of this study are worth noting as well. The cross-sectional design limits the ability to establish causal relationships between LLIN coverage, utilization, and malaria incidence, as it captures only a snapshot of the situation during data collection. Moreover, while disparities in LLIN access and utilization among the FDMN population were highlighted, the study did not explore the underlying sociocultural or logistical factors contributing to these gaps. Finally, the geographic coverage, while extensive, may not be entirely generalizable to other malaria-endemic areas of Bangladesh. Despite these limitations, the study provides valuable insights into LLIN coverage and utilization in Bangladesh.

## Conclusion

In conclusion, these studies offer significant insights for the design and implementation of targeted malaria control interventions in Bangladesh. The findings highlight the necessity for strategies tailored to specific regions, considering demographic differences and variations in LLIN usage. Public health initiatives must tackle these complex challenges to improve the effectiveness of LLIN programs and support the overarching objective of malaria prevention.

## Supporting information

**S1 File. The structured questionnaire.**

**S1 Checklist. The STROBE statement checklist for cross sectional study**

## Author Contributions

Conceptualization, M.S.H., A.K.N., N.-E.N.F. and M.S.A.; methodology, M.S.H., A.K.N, C.W.P, N.-E.N.F. and M.S.A.; software, M.S.H.; validation, M.S.H., A.K.N. and N.-E.N.F.; formal analysis, M.S.H. and M.S.A.; investigation, M.S.H., A.K.N., C.W.P, N.-E.N.F., A.H., M.M.R. (Md. Mushfiqur Rahman), M.M.R. (Md. Musiqure Rahman), S.K.D., S.I., M.N.I., A.T.M.R.H.B, M.A.I. and M.S.A; data curation, M.S.H., A.K.N., C.W.P. and N.-E.N.F.; writing—original draft preparation, M.S.H. and M.S.A.; writing—review and editing, M.S.H., A.K.N., N.-E.N.F., C.W.P., A.H., S.I., M.M.R. (Md. Mushfiqur Rahman), M.M.R. (Md. Musiqure Rahman), M.N.I, S.K.D., A.T.M.R.H.B, M.N.I. (Md. Nazmul Islam), M.A.I. and M.S.A.; visualisation, M.S.H. and A.H.; supervision, S.I., M.M.R. (Md. Mushfiqur Rahman), M.M.R. (Md. Musiqure Rahman), M.N.I, S.K.D., A.T.M.R.H.B, M.N.I. (Md. Nazmul Islam), M.A.I. and M.S.A.; funding acquisition, M.S.H., M.A.I. and M.S.A. All authors have read and agreed to the published version of the manuscript.

## Institutional Review Board Statement

The study was conducted in accordance with the Declaration of Helsinki, and approved by the Institutional Review Board of icddr,b (Protocol no: PR-20097 and date of approval: 16 April 2023).

## Informed Consent Statement

Informed consent was obtained from all subjects involved in the study.

## Data Availability Statement

The data presented in this study are available from the corresponding author (M.S.A.) if requested reasonably.

## Acknowledgments

We express our gratitude to all the individuals who participated in this study and to the dedicated personnel involved. This research study was funded by BRAC through the Global Fund to Fight AIDS, Tuberculosis, and Malaria (GFATM). The icddr,b acknowledges with gratitude the commitment of BRAC and the Global Fund to Fight AIDS, Tuberculosis, and Malaria (GFATM) to its research efforts. The icddr,b is also grateful to the Governments of Bangladesh and Canada for providing core/unrestricted support.

## Reference

1. Birhanu Z, Yihdego YY-e, Yewhalaw D. Caretakers’ understanding of malaria, use of insecticide treated net and care seeking-behavior for febrile illness of their children in Ethiopia. BMC infectious diseases. 2017;17:1–16.

2. Organization WH. World malaria report 2023: World Health Organization; 2023.

3. DGHS. National strategic plan for malaria elimination and prevention of re-establishment of malaria transmission in Bangladesh 2024-2030. Ministry of Health and Family Welfare,Government of the People’s Republic of Bangladesh: Dhaka, Bangladesh.

4. Alam MS, Kabir MM, Hossain MS, Naher S, Ferdous NE, Khan WA, et al. Reduction in malaria prevalence and increase in malaria awareness in endemic districts of Bangladesh. Malar J. 2016;15(1):552.

5. Amin MA. Bangladesh is gunning for zero malaria deaths by 2027 2024 [Available from: https://www.gavi.org/vaccineswork/bangladesh-gunning-zero-malaria-deaths-2027#:∼:text=level%20health%20worker.-,Both%20Bangladesh’s%20malaria%20case%2Dcount%20and%20its%20malaria%20fatality%20rate,the%20space%20of%20a%20year.

6. DGHS. Health Bulletin 2023. Ministry of Health and Family Welfare,Government of the People’s Republic of Bangladesh: Dhaka, Bangladesh.; 2024.

7. Hossain MS, Matin MA, Ferdous N-EN, Hasan A, Sazed SA, Neogi AK, et al. Adherence to Anti-Malarial Treatment in Malaria Endemic Areas of Bangladesh. Pathogens. 2023;12(12):1392.

8. A Record Review on the Health Status of Rohingya Refugees in Bangladesh. [Internet]. [cited 2024 Nov 28] [28 Nov 2024]. Available from: https://www.cureus.com/articles/34588-a-record-review-on-the-health-status-of-rohingya-refugees-in-bangladesh.

9. A Record Review on the Health Status of Rohingya Refugees in Bangladesh [Internet]. [cited 2022 Apr 10]. [Available from: https://www.cureus.com/articles/34588-a-record-review-on-the-health-status-of-rohingya-refugees-in-bangladesh.

10. The Rohingya crisis: A health situation analysis of refugee camps in Bangladesh [Internet]. [cited 2022 Apr 10]. [Available from: https://www.orfonline.org/research/the-rohingya-crisis-a-health-situation-analysis-of-refugee-camps-in-bangladesh-53011/.

11. Jeffries R, Abdi H, Ali M, Bhuiyan ATMRH, El Shazly M, Harlass S, et al. The health response to the Rohingya refugee crisis post August 2017: Reflections from two years of health sector coordination in Cox’s Bazar, Bangladesh. Plos one. 2021;16(6):e0253013.

12. Haque U, Overgaard HJ, Clements AC, Norris DE, Islam N, Karim J, et al. Malaria burden and control in Bangladesh and prospects for elimination: an epidemiological and economic assessment. The Lancet Global Health. 2014;2(2):e98–e105.

13. Haque U, Ahmed SM, Hossain S, Huda M, Hossain A, Alam MS, et al. Malaria prevalence in endemic districts of Bangladesh. PloS one. 2009;4(8):e6737.

14. Kabir MM, Naher S, Islam A, Karim A, Rasid MH-O, Laskar SI. Vector control using LLIN/ITN: reduction of malaria morbidity in Bangladesh. Malaria journal. 2014;13(Suppl 1):P47.

15. Mohammad Sharif Hossain CSP, Mohammad Shafiul Alam, Rashidul Haque, Wasif Ali Khan, Md. Akramul Islam, Shayla Islam, Md. Ekramul Haque, Md. Nazmul Islam Utilization of long lasting Insecticidal Nets (LLINs), knowledge & practices on malaria among the people in malaria endemic districts of Bangladesh and Forcibly Displaced Myanmar Nationals (FDMN) Study Report 2021. icddr,b; BRAC and DGHS, Ministry of Health and Family Welfare,Government of the People’s Republic of Bangladesh: Dhaka, Bangladesh.; 2022.

16. Ahmed SM, Hossain S, Kabir MM, Roy S. Free distribution of insecticidal bed nets improves possession and preferential use by households and is equitable: findings from two cross-sectional surveys in thirteen malaria endemic districts of Bangladesh. Malaria journal. 2011;10:1–9.

17. Khanam F, Hossain MB, Chowdhury TR, Rahman MS, Kabir M, Naher S, et al. Exploring the gap between coverage, access, and utilization of long-lasting insecticide-treated nets (LLINs) among the households of malaria endemic districts in Bangladesh. Malaria Journal. 2018;17:1–12.

18. Raghavendra K, Chourasia MK, Swain DK, Bhatt RM, Uragayala S, Dutta G, et al. Monitoring of long-lasting insecticidal nets (LLINs) coverage versus utilization: a community-based survey in malaria endemic villages of Central India. Malaria Journal. 2017;16:1–8.

19. Wanzira H, Eganyu T, Mulebeke R, Bukenya F, Echodu D, Adoke Y. Long lasting insecticidal bed nets ownership, access and use in a high malaria transmission setting before and after a mass distribution campaign in Uganda. PloS one. 2018;13(1):e0191191.

20. Joshi A, Banjara M. Malaria related knowledge, practices and behaviour of people in Nepal. Journal of Vector Borne Diseases. 2008;45(1):44.

21. Diema Konlan K, Japiong M, Dodam Konlan K, Afaya A, Salia SM, Kombat JM. Utilization of insecticide treated bed nets (ITNs) among caregivers of children under five years in the Ho municipality. Interdisciplinary perspectives on infectious diseases. 2019;2019(1):3693450.

22. Kanyangarara M, Hamapumbu H, Mamini E, Lupiya J, Stevenson JC, Mharakurwa S, et al. Malaria knowledge and bed net use in three transmission settings in southern Africa. Malaria Journal. 2018;17:1–12.

23. Kulkarni MA, Eng JV, Desrochers RE, Cotte AH, Goodson JL, Johnston A, et al. Contribution of integrated campaign distribution of long-lasting insecticidal nets to coverage of target groups and total populations in malaria-endemic areas in Madagascar. The American journal of tropical medicine and hygiene. 2010;82(3):420.

24. Organization WH. Achieving universal coverage with long-lasting insecticidal nets in malaria control. Global Malaria Programme Geneva: WHO. 2014.

25. Tassew A, Hopkins R, Deressa W. Factors influencing the ownership and utilization of long-lasting insecticidal nets for malaria prevention in Ethiopia. Malaria journal. 2017;16:1–9.

26. Ng’ang’a PN, Aduogo P, Mutero CM. Long lasting insecticidal mosquito nets (LLINs) ownership, use and coverage following mass distribution campaign in Lake Victoria basin, Western Kenya. BMC Public Health. 2021;21(1):1046.

27. Pooseesod K, Parker DM, Meemon N, Lawpoolsri S, Singhasivanon P, Sattabongkot J, et al. Ownership and utilization of bed nets and reasons for use or non-use of bed nets among community members at risk of malaria along the Thai-Myanmar border. Malaria journal. 2021;20:1–12.

28. Birhanu Z, Abebe L, Sudhakar M, Dissanayake G, Yihdego Y, Alemayehu G, et al. Access to and use gaps of insecticide-treated nets among communities in Jimma Zone, southwestern Ethiopia: baseline results from malaria education interventions. BMC Public Health. 2015;15:1–11.

29. Kilian A, Koenker H, Baba E, Onyefunafoa EO, Selby RA, Lokko K, et al. Universal coverage with insecticide-treated nets–applying the revised indicators for ownership and use to the Nigeria 2010 malaria indicator survey data. Malaria journal. 2013;12:1–12.

30. Organization WH. Recommendations for Achieving Universal Coverage with Long-Lasting Insecticidal Nets in Malaria Control. Geneva: World Health Organization; 2014.

31. Seyoum D, Speybroeck N, Duchateau L, Brandt P, Rosas-Aguirre A. Long-lasting insecticide net ownership, access and use in Southwest Ethiopia: a community-based cross-sectional study. International journal of environmental research and public health. 2017;14(11):1312.

32. Adaji J, Gabriel OE. Access and Usage of Long Lasting Insecticidal Nets (LLIN) in rural Communities of Benue State, Nigeria. Health Science Journal. 2019;13(1):1–4.

33. Ouedraogo LT, Ouedraogo I, Yameogo A, Ouedraogo V. Determinants of long-lasting insecticidal net use in Burkina Faso after a mass distribution in the Diebougou health district. Revue d’epidemiologie et de sante publique. 2013;61(2):121–7.

34. Negash K, Haileselassie B, Tasew A, Ahmed Y, Getachew M. Ownership and utilization of long-lasting insecticide-treated bed nets in Afar, northeast Ethiopia: a cross-sectional study. The Pan African Medical Journal. 2012;13(Suppl 1).

35. Bisong CE, Dongmo CM. Utilization of malaria prevention methods by pregnant women in Yaounde. Pan African Medical Journal. 2013;15(1).

36. Jambulingam P, Gunasekaran K, Sahu S, Vijayakumar T. Insecticide treated mosquito nets for malaria control in India-experience from a tribal area on operational feasibility and uptake. Memórias do Instituto Oswaldo Cruz. 2008;103:165–71.

37. Stebbins RC, Emch M, Meshnick SR. The effectiveness of community bed net use on malaria parasitemia among children less than 5 years old in Liberia. The American Journal of Tropical Medicine and Hygiene. 2018;98(3):660.

38. Githinji S, Herbst S, Kistemann T, Noor AM. Mosquito nets in a rural area of Western Kenya: ownership, use and quality. Malaria journal. 2010;9:1–6.

39. Kateera F, Ingabire CM, Hakizimana E, Rulisa A, Karinda P, Grobusch MP, et al. Long-lasting insecticidal net source, ownership and use in the context of universal coverage: a household survey in eastern Rwanda. Malaria journal. 2015;14:1–10.

